# Associations of Body Mass Index on worsening of heart failure and mortality in patients with heart failure and reduced left ventricular ejection fraction: A 10-year follow-up study (A NorthStar Substudy)

**DOI:** 10.1101/2025.05.05.25327040

**Authors:** Morten Malmborg, Mohamed El-Chouli, Camilla Fuchs Andersen, Mariam Elmegaard, Caroline Garred, Deewa Zahir, Jawad H Butt, Daniel M Christensen, Nina Nouhravesh, Emil Fosbøl, Lars Videbæk, Lars Køber, Finn Gustafsson, Morten Schou

## Abstract

**Background:** Obesity is common in heart failure with reduced ejection fraction (HFrEF). As anti-obesity treatments advance, understanding how body mass index (BMI) affects outcomes in HFrEF is increasingly important.

**Objective:** To examine whether a BMI >27 kg/m² is linked to higher risks of all-cause mortality, cardiovascular death, and heart failure (HF) hospitalization in HFrEF patients.

**Methods:** This study included 1,017 clinically stable, medically optimized HFrEF patients from the NorthStar study (2005–2009), followed through 2023 using Danish registries. Outcomes were assessed with Cox models adjusted for prognostic factors. The primary endpoint was all-cause mortality; secondary endpoints included cardiovascular death, HF hospitalization, and a composite of mortality or hospitalization. Subgroup analyses compared BMI categories (<24, 24–27, >27 kg/m²).

**Results:** Patients with BMI >27 had more diabetes (27.8% vs. 17.7%) and lower NT-proBNP (median 776 vs. 1,163 pg/mL) than those with BMI 24–27, with similar HF etiology. Over a median 8.8 years, 821 patients (80.7%) died, including 444 cardiovascular deaths, and 740 (72.8%) were hospitalized for HF. A BMI of 35 vs. 27 was associated with non-significant increased all-cause mortality (HR 1.18, 95% CI 0.94–1.48) but significantly higher cardiovascular mortality (HR 1.42, 95% CI 1.05–1.92), HF hospitalization (HR 1.33, 95% CI 1.05–1.67), and composite outcome (HR 1.30, 95% CI 1.06–1.60). Subgroup analysis showed higher mortality with BMI >27 vs. 24–27 in ischemic cardiomyopathy (HR 1.31, 95% CI 1.05–1.64), but not in non-ischemic (HR 0.86, 95% CI 0.66–1.12), interaction p=0.015.

**Conclusion:** Among HFrEF patients—especially those with ischemic cardiomyopathy—BMI >27 is associated with worse outcomes, challenging the “obesity-survival paradox” and highlighting the importance of effective weight management.

## Introduction

Heart failure (HF) with reduced ejection fraction (HFrEF) remains a major global public health issue, characterized by high rates of morbidity and mortality.^1,2^ Obesity is a common comorbidity in patients with HFrEF, further complicating disease management and prognosis.^3^ With the recent emergence of effective anti-obesity drugs, a clearer understanding of the relationship between body mass index (BMI) and clinical outcomes in HFrEF has become increasingly important.

The association between BMI and outcomes in patients with HFrEF has long been a subject of debate. While elevated BMI is conventionally linked to increased cardiovascular risk,^4–6^ several studies suggest an “obesity paradox”, where higher BMI may confer a survival benefit in patients with chronic diseases, including HF.^7–11^ However, emerging evidence challenges this paradox.^12^ For example, a post-hoc analysis of the DANISH trial (The Danish Study to Assess the Efficacy of Implantable Cardioverter Defibrillators in Patients With Nonischemic Systolic Heart Failure on Mortality) that included long-term follow-up demonstrated a clear association between greater adiposity and increased mortality.^13^ Notably, the DANISH trial only included patients with non-ischemic cardiomyopathy, limiting its ability to assess differences based on HF etiology.

Furthermore, despite natriuretic peptide levels being one of the most significant prognostic factors in HFrEF, few studies examining the relationship between BMI (or other anthropometric measures) and outcomes have accounted for this variable.

This study aims to assess whether an elevated BMI is linked to higher rates of all-cause mortality, cardiovascular mortality, and HF hospitalizations in patients with HFrEF, considering HF etiology.

## Methods

### Study design

This study is a post-hoc, registry-based follow-up of patients initially enrolled in the NorthStar Study (1,017 patients in total). The NorthStar Study was an investigator-initiated, multicenter, randomized, open-label trial with blinded endpoint evaluation (PROBE), which has been described in detail previously.^14,15^ In brief, patients with HFrEF were educated on self-care and optimized for medical therapy in accordance with contemporary HF guidelines at specialized HF clinics. Following this, participants were randomized to either continued follow-up in a specialized HF clinic or discharge to primary care. During the trial, patients were followed for a median of 2.5 years, with a maximum follow-up period of five years. The intervention demonstrated no significant impact on the primary composite outcome (death from any cause or hospitalization for a protocol-specified cardiovascular cause), cardiovascular hospitalization, or all-cause mortality.^14^

### Data sources

In Denmark, all residents are assigned a unique and permanent civil registration number at birth or upon immigration, allowing for individual-level linkage between nationwide registries and the electronic case report forms from the NorthStar study. Baseline data, including demographic characteristics, clinical biochemistry, left ventricular ejection fraction, and N-terminal pro-b-type natriuretic peptide (NT-proBNP) levels, were sourced from the trial’s electronic case report form.

For this extended follow-up analysis, additional data were obtained from the following sources:

1. The Danish Civil Registration System (information on emigration and vital status),
2. The Danish National Patient Registry (hospital discharge diagnoses coded using the International Classification of Diseases (ICD)-10 since 1994), and
3. The Danish National Causes of Death Registry (date and underlying cause of death based on death certificates).

All registries used in this study have been validated previously.^16–18^

### Study population

Patients with HFrEF (defined as a left ventricular ejection fraction <0.45) were enrolled between 2005 and 2009 from 18 of the 40 public HF clinics in Denmark, based on previously described inclusion and exclusion criteria.^14,15^ The present analysis included all patients with a recorded BMI (n = 1,017). At baseline, patients received education on self-care and HF management and were on optimized medical therapy.

Up-titration of renin-angiotensin-system inhibitors, beta-blockers, and mineralocorticoid receptor antagonists was conducted in accordance with international guidelines, with device implantation performed when clinically indicated.^19^ At the time of inclusion into the NorthStar study, an ejection fraction <0.45 was the threshold for diagnosing HFrEF in Denmark.

### Study outcomes

The primary outcome of the study was all-cause mortality. Secondary outcomes included cardiovascular (CV) mortality (ICD-10 codes I00–I99), hospitalization for HF (defined as an overnight stay with one of the following HF admission codes: ICD-10 codes I110, I42, I50, J819), and the composite outcome of all-cause mortality or HF hospitalization, whichever occurred first. HF admission diagnoses have been validated with a positive predictive value of 81%.^20^

### Statistical analyses

All individuals were followed from the date of enrollment until the earliest occurrence of an event of interest, death, emigration, or 31 December 2022.

Baseline characteristics were summarized as frequencies and percentages for categorical variables and medians with interquartile ranges for continuous variables, categorized by BMI groups (<24, 24–27, >27 kg/m²).

The primary analysis examined the association between BMI, treated as a continuous variable, and the outcomes of interest using restricted cubic splines in Cox proportional-hazards models. Hazard ratios (HR) with 95% confidence intervals (CI) were reported, using a BMI of 27 kg/m² as the reference – a cut-off used in some randomized clinical trials.^21,22^ Models were adjusted for known key prognostic factors: sex, age (<70, ≥70 years), estimated glomerular filtration rate (≤60, >60), NT-proBNP level (<1000, ≥1000), New York Heart Association class (1–2, 3), diabetes (yes, no), atrial fibrillation (yes, no), HF etiology (ischemic, non-ischemic), left ventricular ejection fraction (≤30, >30), use of mineralocorticoid receptor antagonists, beta-blockers, and Renin-angiotensin-aldosterone system inhibitors.

Subgroup analyses based on the aforementioned, adjusted Cox models were conducted for the primary outcome to assess potential interactions between BMI categories and prognostic factors. Two comparisons were evaluated: (1) BMI <24 versus 24–27 (reference) and (2) BMI 24–27 (reference) versus >27.

A supplementary analysis examined the adjusted association between BMI as a continuous variable and outcomes, stratified by HF etiology. Patients with ischemic cardiomyopathy and a BMI of 27 kg/m² served as the reference group.

All statistical analyses were two-sided with a significance level of 5% and performed using R version 4.4.1.^23^

### Ethics

The trial was approved by the Danish Ethics Committee (KF 01/2724936) and the extended follow-up was approved by the Danish Data Protection Agency (Approval number P-2019-348).

## Results

### Population characteristics

At enrollment, the median age of the population was 69 years, 75% were men, and the median BMI was 26.3 kg/m² (Table 1). Treatment rates with RASi and BB were high, while only 31% were receiving an MRA at baseline (Table 1). None were treated with sodium-glucose transport protein 2 inhibitors or glucagon-like peptide-1 receptor analogues as reflecting the inclusion period. Compared to patients with a BMI of 24–27 kg/m², those with a BMI >27 kg/m² had a higher prevalence of diabetes (27.8% vs. 17.7%), similar rates of ischemic HF etiology (57.5% vs. 58.7%), lower NT-proBNP levels (median 776 vs. 1,163 pg/mL), and a higher use of mineralocorticoid receptor antagonists (35.7% vs. 27.5%). Patients with a BMI <24 kg/m², were older (median 73 vs. 69 years), included fewer men (65.7% vs. 80.4%), had a lower prevalence of ischemic cardiomyopathy (54.7% vs. 58.7%) and diabetes (8.1% vs. 17.7%), but had higher NT-proBNP levels (median 1,421 vs. 1,163 pg/mL).

**Table 1:** Population characteristics at enrollment.

| Variable | BMI <24 | BMI 24-27 | BMI >27 | Total |
| --- | --- | --- | --- | --- |
| **N** | 309 | 327 | 471 | 1107 |
| **BMI (IQR)** | 22.5 [20.9, 23.2] | 25.5 [24.7, 26.3] | 30.3 [28.4, 32.8] | 26.3 [23.8, 29.6] |
| **Age (IQR)** | 73 [65, 79] | 69 [62, 77] | 68 [61, 74] | 69 [62, 76] |
| **Men** | 203 (65.7) | 263 (80.4) | 368 (78.1) | 834 (75.3) |
| **HF etiology (Ischemic)** | 169 (54.7) | 192 (58.7) | 271 (57.5) | 632 (57.1) |
| **Stroke or TCI** | 47 (15.2) | 34 (10.4) | 55 (11.7) | 136 (12.3) |
| **PAD** | 29 (9.4) | 23 (7.1) | 49 (10.4) | 101 (9.2) |
| **Atrial fibrillation** | 104 (33.7) | 107 (32.7) | 168 (35.7) | 379 (34.2) |
| **Diabetes** | 25 (8.1) | 58 (17.7) | 131 (27.8) | 214 (19.3) |
| **COPD** | 55 (17.8) | 38 (11.6) | 72 (15.3) | 165 (14.9) |
| **LVEF (IQR)** | 30 [25.0, 37.8] | 30 [25, 37] | 34 [25, 40] | 30 [25, 40] |
| **NT-proBNP (IQR)** | 1,421 [617, 2,702] | 1,163 [473.5, 2,126.5] | 776 [268, 1,710] | 1,126 [382.5, 2,080.0] |
| **eGFR (IQR)** | 64.4 [50.9, 82.0] | 66.7 [52.8, 80.9] | 67.8 [53.0, 83.4] | 66.7 [52.2, 82.6] |
| **NYHA 1-2** | 277 (89.6) | 295 (90.2) | 413 (87.7) | 985 (89.0) |
| **BB** | 253 (81.9) | 276 (84.4) | 407 (86.4) | 936 (84.6) |
| **RAASi** | 265 (85.8) | 289 (88.4) | 399 (84.7) | 953 (86.1) |
| **MRA** | 84 (27.2) | 90 (27.5) | 168 (35.7) | 342 (30.9) |
| **ICD** | 29 (9.4) | 37 (11.3) | 44 (9.4) | 110 (10.0) |
| **CRT** | 8 (2.6) | 17 (5.2) | 19 (4.1) | 44 (4.0) |

**Abbreviations:**
BMI, body mass index; IQR, interquartile range; HF, heart failure; TCI, transitory cerebral ischemia; PAD, peripheral arterial disease; COPD, chronic obstructive pulmonary disorder; LVEF, left ventricular ejection fraction; NT-proBNP, amino-terminal-pro-brain-natriuretic-peptide; eGFR, estimated glomerular filtration rate; NYHA class, New York Heart Association; BB, beta-blocker; RAASi, Renin-angiotensin-aldosterone system inhibitors; MRA, mineralocorticoid receptor antagonists; ICD, implantable cardiac defibrillator; CRT, cardiac resynchronization therapy.

### Rates of primary and secondary outcomes

During follow-up, 821 patients (80.7%) died, of which 444 were classified as cardiovascular deaths. A total of 740 patients (72.8%) were hospitalized for HF and 961 composite events (HF hospitalization or all-cause death) were observed.

Adjusted rates of all-cause mortality increased with higher BMI above 27, though the association was not statistically significant (BMI 35 vs 27: HR 1.18 [95% CI: 0.94–1.48]; Figure 1). Similar patterns were observed for secondary outcomes, including cardiovascular death (BMI 35 vs 27: HR 1.42 [1.05–1.92]), HF hospitalization (BMI 35 vs 27: HR 1.33 [1.05–1.67]), and the composite outcome (BMI 35 vs 27: HR 1.30 [1.06–1.60]). Subgroup analysis revealed significantly higher mortality rates for BMI >27 compared to 24–27 in patients with ischemic cardiomyopathy (HR 1.31 [1.05–1.64]), but not in those with non-ischemic cardiomyopathy (HR 0.86 [0.66–1.12]; p for interaction = 0.015; Figure 2).

**Figure 1.**
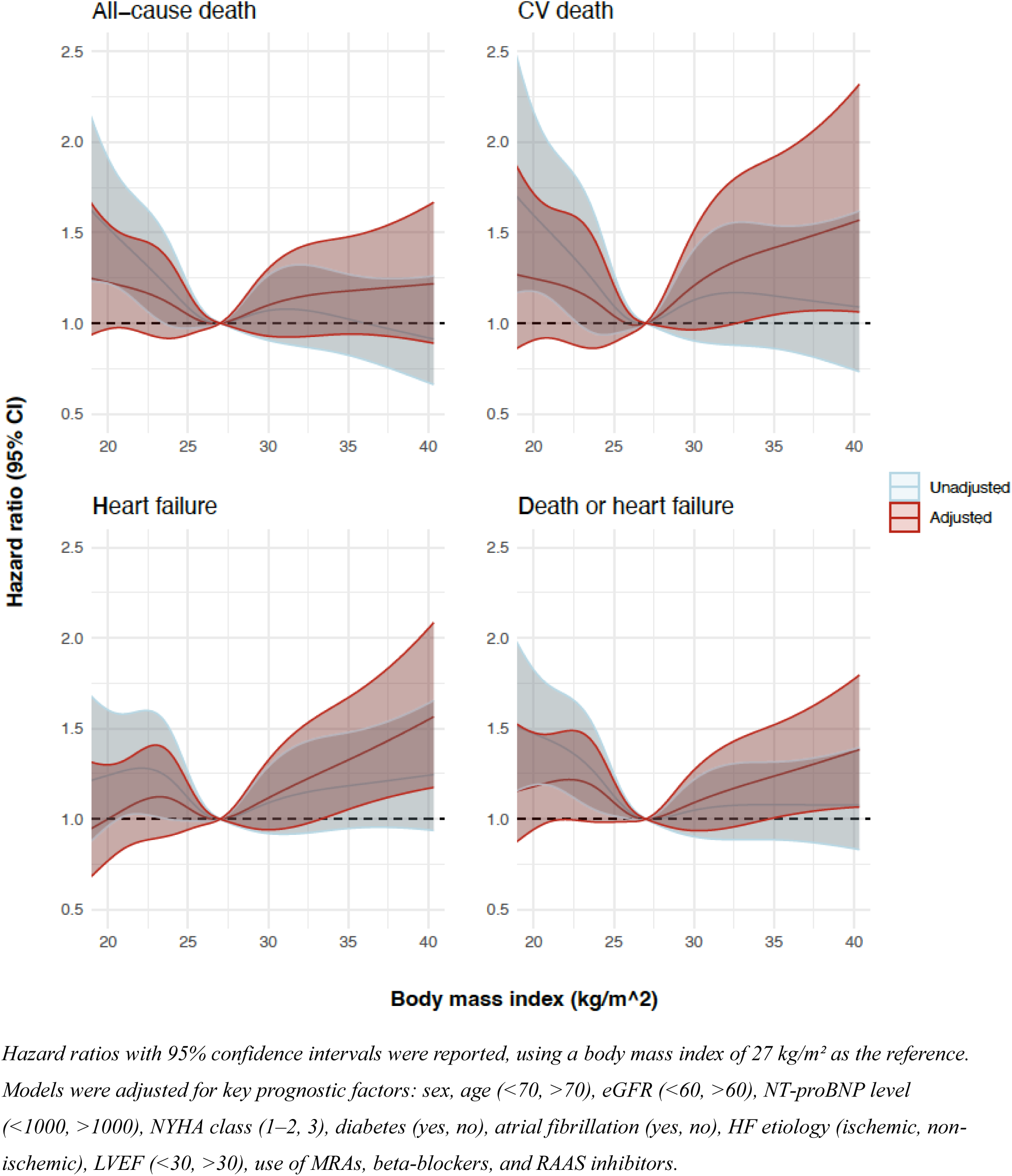
Associations between body mass index, treated as a continuous variable, and the primary and secondary outcomes using restricted cubic splines in Cox proportional-hazards models.

**Figure 2.**
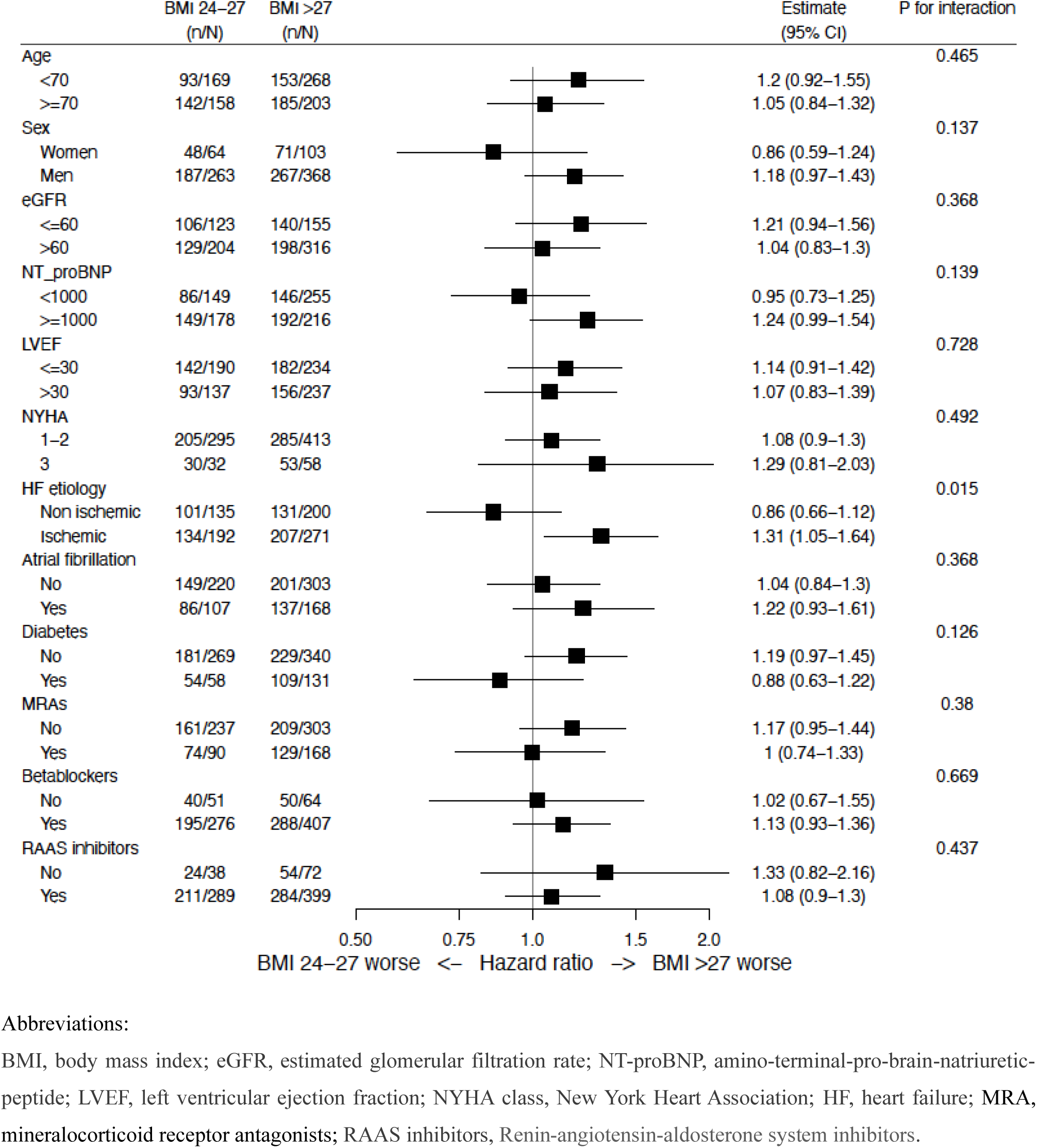
Subgroup analyses for the primary outcome based on interactions between prognostic factors and body mass index groups >27 vs. 24–27 kg/m² (reference).

Adjusted rates for all-cause mortality increased non-significantly with BMI <27 (e.g., BMI 20 vs 27: HR 1.23 [0.97–1.55]; Figure 2). Similar trends were seen for cardiovascular death (BMI 20 vs 27:

HR 1.25 [0.90–1.72]) and the composite outcome (BMI 20 vs 27: HR 1.17 [0.94–1.46]), but not for HF hospitalization (BMI 20 vs 27: HR 1.00 [0.77–1.30]). A significant interaction for HF etiology was observed (p for interaction = 0.002), where patients with ischemic cardiomyopathy, but not non-ischemic cardiomyopathy, had higher all-cause mortality rates with BMI <24 compared to 24–27 (Figure 3). Additionally, male sex and diabetes were associated with higher all-cause mortality rates in patients with a BMI <24 (p for interaction = 0.018 and 0.036, respectively).

**Figure 3.**
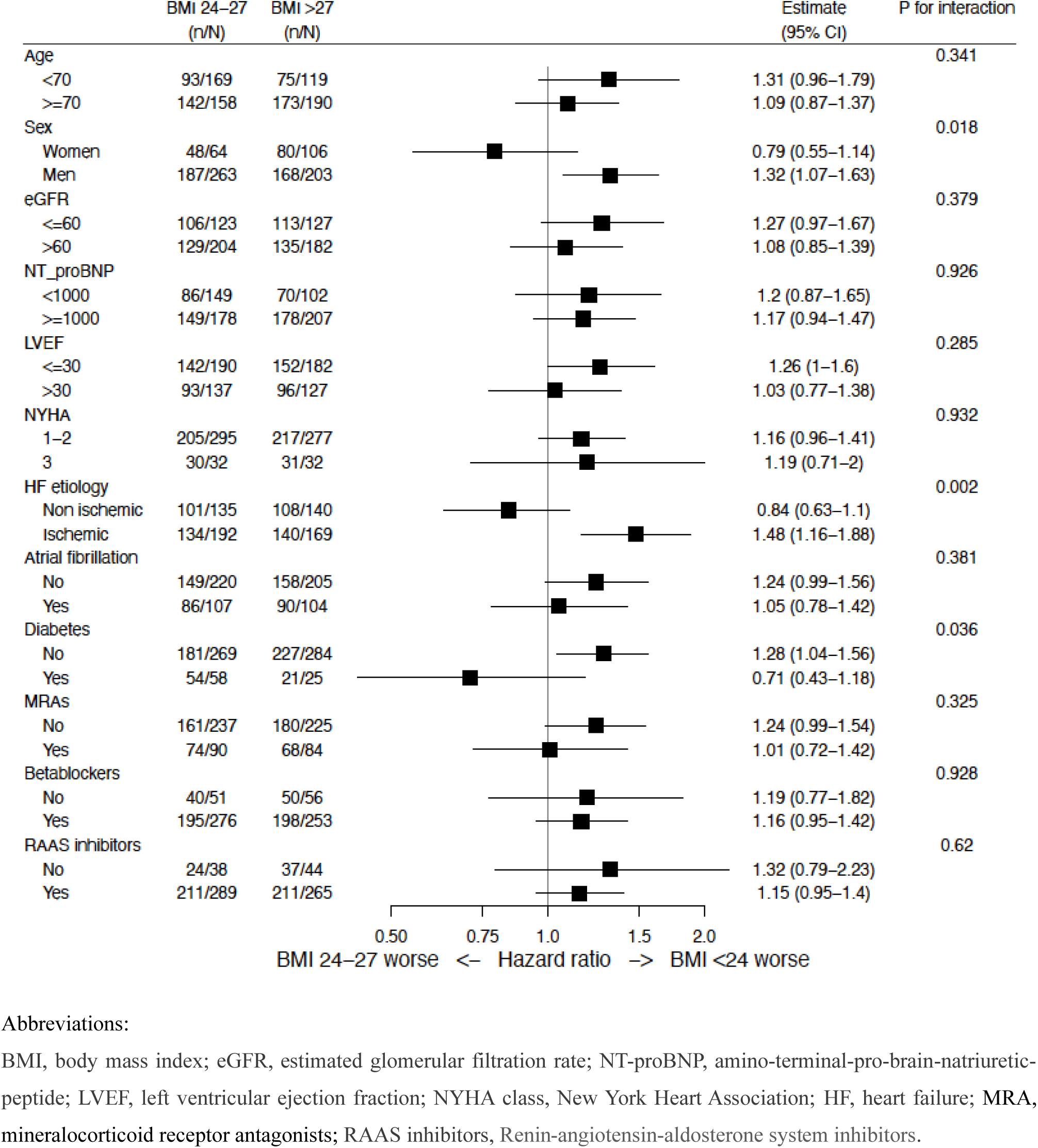
Subgroup analyses for the primary outcome based on interactions between prognostic factors and body mass index groups <24 vs. 24–27 (reference).

### Supplementary analyses

In supplementary analyses, adjusted rates of primary and secondary outcomes, especially cardiovascular death, increased with BMI >27 kg/m² (Figure 4). However, these associations were predominantly observed in HF patients with ischemic cardiomyopathy.

**Figure 4.**
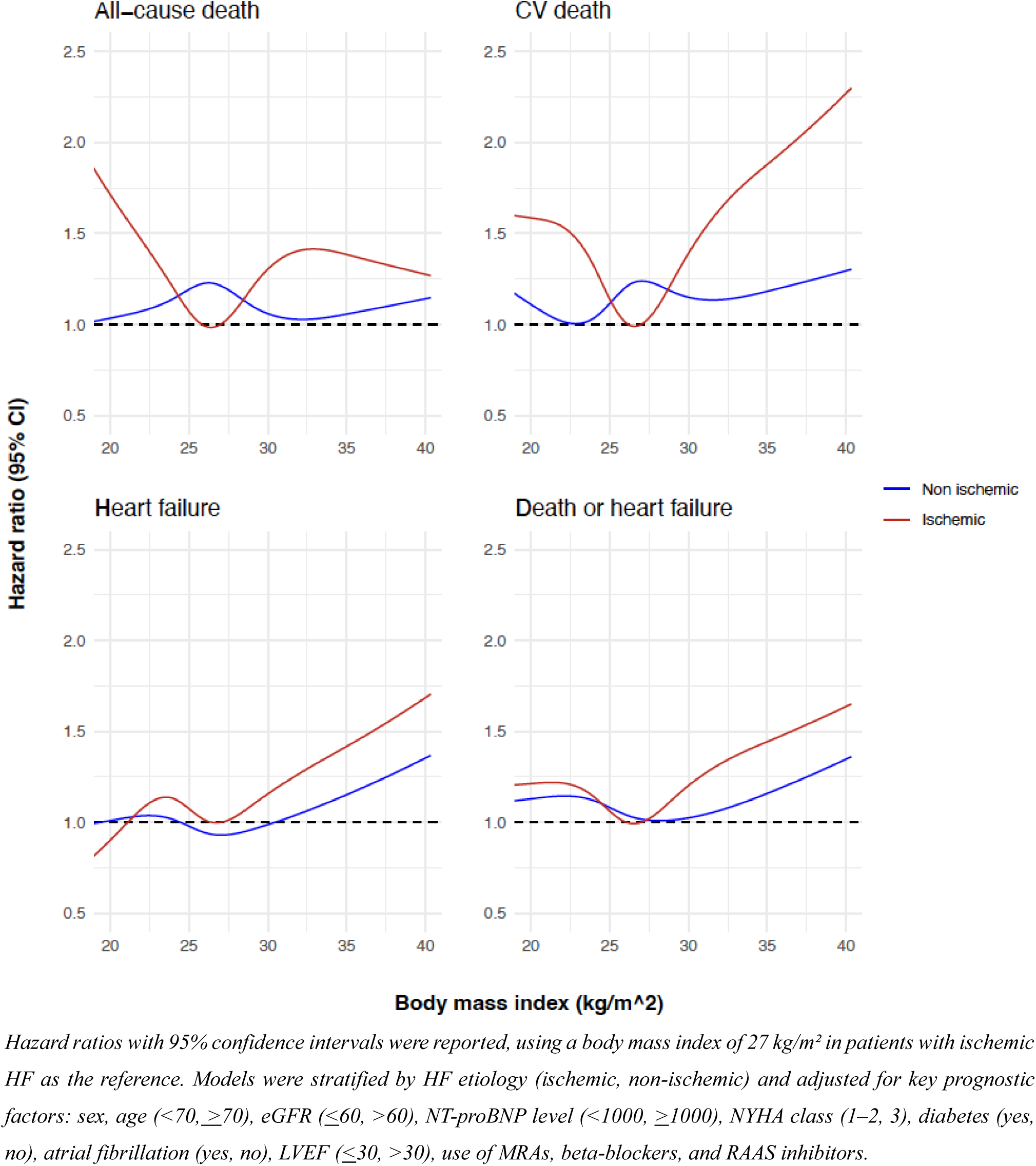
Associations between body mass index treated as a continuous variable, and the primary and secondary outcomes stratified by heart failure etiology using restricted cubic splines in Cox proportional-hazards models.

## Discussion

This study utilized data from the NorthStar study to evaluate the association between BMI and clinical outcomes in patients with HFrEF. The key findings were that a BMI >27 kg/m² was associated with increased rates all-cause mortality, CV mortality, and HF hospitalization. These results challenge the “obesity-survival paradox,” which posits that elevated BMI might offer survival advantages in chronic diseases such as HF. Importantly, the adverse impact of BMI >27 kg/m², was more pronounced in HF patients with ischemic cardiomyopathy, while non-ischemic HF showed no significant association, emphasizing the heterogeneity in HF subtypes.

While several studies suggest an “obesity paradox”,^7–11^ our results align with emerging evidence challenging this notion. For instance, the PARADIGM-HF trial did observe better outcomes among HF patients with elevated BMI, but not when adjusting for NT-proBNP level, suggesting that obesity may not be protective.^12^ However, it also did not demonstrate an increased mortality risk associated with higher BMI. The relatively short follow-up period of PARADIGM-HF (median follow-up of 27 months) likely limited its ability to fully capture the long-term impact of obesity on mortality, which may only become apparent with extended follow-up. Consistent with our findings, the DANISH trial identified an association between obesity and higher mortality, including cardiovascular mortality.^13^ These similarities strengthen the robustness of our conclusion that

BMI >27 kg/m², may be a risk factor rather than a protective marker in this context. However, the DANISH trial was limited to patients with non-ischemic cardiomyopathy and included a younger cohort, making direct comparisons challenging.

To our knowledge, this is the first study to identify a significant interaction between BMI and HF etiology, demonstrating an increased mortality risk in patients with ischemic cardiomyopathy but not in those with non-ischemic cardiomyopathy. While no previous study has reported higher mortality in ischemic cardiomyopathy patients with elevated BMI, Gentile et al. observed improved survival rates among overweight and obese HF patients with non-ischemic cardiomyopathy, but not in their ischemic counterparts.^24^ Furthermore, recent post-hoc results from the SELECT trial demonstrated that semaglutide significantly reduced CV events and improved HF outcomes in HF patients with atherosclerotic cardiovascular disease and a BMI of >27, irrespective of HF subtype.^21^ These results lend additional support to our findings that emphasize the importance of weight management in HF patients with elevated BMI, especially in ischemic HF subtypes.

Mechanistically, obesity-related metabolic disturbances, such as insulin resistance, systemic inflammation, and altered adipokine signaling, may amplify ischemic injury in cardiomyocytes, exacerbating disease progression in this subgroup.^25^ Conversely, the lack of a similar relationship in non-ischemic cardiomyopathy might reflect differing underlying pathophysiological mechanisms, such as those driven by genetic cardiomyopathies, which are less influenced by obesity.

Interestingly, our data also revealed worse outcomes in patients with a BMI <24 kg/m², particularly in ischemic cardiomyopathy. This observation could reflect advanced disease or cachexia, which are well-documented predictors of poor prognosis in HF. The adverse impact of low BMI is consistent with prior studies showing that frailty and muscle wasting, rather than the lower fat mass itself, drive mortality risk in HF.^26,27^

### Clinical implications

Given the rising prevalence of obesity and the availability of effective anti-obesity therapies, our findings may have significant implications for the management of HFrEF patients. The observed association between elevated BMI and adverse outcomes supports an important role for careful weight management in this population. This may be particularly relevant for patients with ischemic cardiomyopathy, where targeted weight reduction strategies could potentially mitigate risk and improve outcomes. However, the nuances in the relationship between BMI and outcomes, including the absence of a possible protective effect in low-BMI patients, highlight the need for an individualized approach.

Emerging therapies such as GLP-1 receptor agonists have shown promise in reducing weight^22,28^ and improving metabolic health in HF patients with preserved ejection fraction.^29,30^ While these therapies were not evaluated in our study, their potential role in mitigating BMI-related risks in HFrEF warrants further investigation.

### Methodological considerations

A major strength of our study is the large, well-characterized cohort of HFrEF patients with extensive follow-up using validated Danish nationwide registries. The rigorous adjustment for confounders and the stratification by HF etiology provide a detailed understanding of the relationship between BMI and the outcomes of interest. However, certain limitations should be noted. First, BMI was used as a surrogate for obesity, which does not account for differences in body composition, such as visceral versus subcutaneous adiposity. Future studies incorporating advanced measures of body composition, such as computer tomography or magnetic resonance imaging, could provide further insights. Second, this was a post-hoc analysis, precluding causal inferences. No adjustment for multiple testing was performed. Third, our study cohort comprised Danish patients, which may limit generalizability to other populations with differing HF etiologies, BMI distributions, or management strategies.

## Conclusion

In patients with HFrEF, a BMI >27 kg/m² was associated with worse outcomes, particularly in ischemic cardiomyopathy, challenging the “obesity-survival paradox.” Conversely, a BMI <24 kg/m² also increased mortality risk, likely reflecting frailty or advanced disease. These findings support individualized weight management strategies in HFrEF, particularly in obese patients with ischemic HF. Future studies should explore the role of body composition and the potential benefits of targeted weight-loss interventions in this population.

## Data Availability

Data obtained through the nationwide registers in Denmark can be made available only through research on Danish servers hosted in highly protected research environments where researchers can be granted access and permission with encrypted person identification. Access to raw data can be gained only through collaboration with the authors or other Danish institutions that already have been granted access. Please contact the first author with any questions regarding data access.

## Acknowledgments

The technical assistance from the staff at the Department of Clinical Chemistry, Frederiksberg University Hospital is deeply acknowledged. M. Malmborg and M. Schou had full access to all the data in the study and took responsibility for the integrity of the data and the accuracy of the data analysis.

